# A County-Level Susceptibility Index and Coronavirus Disease 2019 Mortality in the United States: A Socioecological Study

**DOI:** 10.1101/2020.07.04.20146084

**Authors:** Sadiya S. Khan, Megan E. McCabe, Amy E. Krefman, Lucia C. Petito, Xiaoyun Yang, Kiarri N. Kershaw, Lindsay Pool, Norrina B. Allen

## Abstract

As of June 2020, the United States (US) has experienced the highest number of deaths related to coronavirus disease 2019 (Covid-19) in the world, but significant geographic heterogeneity exists at the county-level. Therefore, we sought to classify counties in the United States across multiple domains utilizing a socioecological framework and examine the association between these county-level groups and Covid-19 mortality. We harmonized and linked county-level sociodemographic, health, and environmental metrics associated with increased susceptibility for Covid-19 mortality. Latent class analysis defined a county-level susceptibility index (CSI) based on these metrics (n=2701 counties). Next, we used linear regression models to estimate the associations of the CSI and Covid-19 deaths per capita and initial mortality doubling time (as of 6/2/20), adjusted for days since first Covid-19 case. We identified 4 groups classified by the CSI with distinct sociodemographic, health, and environmental profiles and widespread geographic dispersion. Covid-19 deaths per capita were significantly higher in the group consisting of rural, vulnerable counties (55.8 [95% CI 50.3-61.2] deaths per 100,000) compared with the group with diverse, urban counties (32.2 [27.3-37.0]) at similar points in the outbreak (76 days since first case). Our findings can inform equitable resource allocation for Covid-19 to allow targeted public health preparedness and response in vulnerable counties.

## INTRODUCTION

Coronavirus disease 2019 (Covid-19) is caused by a novel severe acute respiratory syndrome coronavirus 2 (SARS-CoV2) and has rapidly emerged into a global pandemic. While the first case was documented to originate in China in November 2019^1,2^, Covid-19 is now widespread across the world^3-6^ and in particular, has a very high burden in the United States (US)^7^. As of June 2020, the US has the highest number of deaths in the world due to Covid-19. While Covid-19 initially affected certain states more heavily in the early period of spread including Washington^8^ and New York^9^, Covid-19 outbreaks have continued to emerge across the US with cases now reported in over 2,600 counties.

To date, clinical and epidemiological data published from China, Italy, and in the US, reported higher rates of symptomatic Covid-19 disease and severity among older adults.^1,8^ In addition to advanced age, history of chronic lung or cardiovascular disease (CVD) are key risk factors of adverse outcomes such as hospitalization and mortality related to severe Covid-19 infection.^10^ Significant race-based disparities^11^ are apparent with 2-fold or higher Covid-19 mortality rates among non-Hispanic blacks compared with non-Hispanic whites^12^ based on reporting from public health departments at the state-level and the Centers for Disease Control (CDC) that mirror and may even exceed disparities noted for non-communicable disease mortality (e.g. CVD and malignancy) in the US.^13,14^

On a population level, mitigating the growing burden related to Covid-19 morbidity and mortality requires identification of communities at higher risk of transmission and mortality.^15,16^ Specifically, identifying counties with similar profiles of scoioecological factors posited to increase community-based Covid-19 transmission and severity of illness will allow an informed public health response for equitable allocation of resources and to mobilize aid in recovery.^17^ It is well-established that wide disparities exist on a county-level in terms of non-communicable disease burden^18^ and life expectancy^19^ that are closely interrelated with sociodemographic,^20^ health^21^, and environmental characteristics^22^. Covid-19 infection susceptibility and mortality may be further amplifying pre-existing place-based health disadvantage. Therefore, we sought to develop a classification scheme integrating information across multiple domains to identify counties at greatest risk to experience a higher burden of morbidity and mortality due to Covid-19 and guide public health responses.

## RESULTS

### County-Level Susceptibility Index (CSI)

Using latent class analysis, we identified 4 classes of counties that represent distinct demographic, socioeconomic, health status, healthcare access, and environmental profiles (Table 1): 1) diverse urban counties with greater social and health assets; 2) rural counties with social and health vulnerabilities; 3) older rural counties with greater social assets; and 4) urban counties with social vulnerabilities. Diverse urban counties had the highest population density, greatest proportion of at least some college education, lowest rates of unemployment and poverty, and fewest absolute years of potential life lost (Supplemental Figure 1). By contrast, vulnerable rural counties had the highest proportion of non-Hispanic black residents, lowest rate of college education, highest rates of unemployment and poverty, and greatest absolute years of potential life lost.

**Table 1.** Demographic, socioeconomic, health status, healthcare access, and environmental factors stratified by the County-Level Susceptibility Index in the United States.

|  | **Diverse Urban**<br>**counties with**<br>**greater social**<br>**assets** | **Rural Counties**<br>**with social and**<br>**medical**<br>**vulnerabilities** | **Older Rural**<br>**counties with**<br>**greater social**<br>**assets** | **Urban Counties**<br>**with social**<br>**vulnerabilities** |
| --- | --- | --- | --- | --- |
| n | 461 | 947 | 712 | 581 |
| **Demographics** |  |  |  |  |
| Population density | 756.84 (2243.99) | 92.35 (447.56) | 29.20 (25.03) | 378.82 (897.47) |
| Proportion male | 0.50 (0.01) | 0.50 (0.03) | 0.51 (0.02) | 0.50 (0.02) |
| Proportion non-Hispanic White | 0.76 (0.16) | 0.69 (0.23) | 0.85 (0.14) | 0.76 (0.17) |
| Proportion non-Hispanic Black | 0.06 (0.07) | 0.17 (0.19) | 0.02 (0.03) | 0.11 (0.11) |
| Proportion Hispanic | 0.11 (0.10) | 0.09 (0.16) | 0.09 (0.13) | 0.09 (0.12) |
| Proportion 60+ years old | 0.23 (0.05) | 0.26 (0.04) | 0.29 (0.06) | 0.24 (0.04) |
| Proportion 80+ years old | 0.04 (0.01) | 0.04 (0.01) | 0.05 (0.01) | 0.04 (0.01) |
| Rural-Urban Continuum Code (%) |  |  |  |  |
| 1 | 212 (46.0) | 36 ( 3.8) | 8 ( 1.1) | 155 (26.7) |
| 2 | 100 (21.7) | 86 ( 9.1) | 28 ( 3.9) | 152 (26.2) |
| 3 | 74 (16.1) | 82 ( 8.7) | 52 ( 7.3) | 122 (21.0) |
| 4 | 31 ( 6.7) | 72 ( 7.6) | 33 ( 4.6) | 77 (13.3) |
| 5 | 14 ( 3.0) | 35 ( 3.7) | 28 ( 3.9) | 12 ( 2.1) |
| 6 | 10 ( 2.2) | 301 (31.8) | 210 (29.5) | 57 ( 9.8) |
| 7 | 15 ( 3.3) | 158 (16.7) | 225 (31.6) | 5 ( 0.9) |
| 8 | 2 ( 0.4) | 83 ( 8.8) | 56 ( 7.9) | 1 ( 0.2) |
| 9 | 3 ( 0.7) | 94 ( 9.9) | 72 (10.1) | 0 ( 0.0) |
| **Socioeconomic Status** |  |  |  |  |
| High school graduation rate | 88.70 (6.25) | 87.97 (7.51) | 89.14 (6.99) | 88.70 (5.34) |
| Percent some college education | 71.09 (7.23) | 48.48 (8.24) | 59.89 (9.31) | 59.30 (7.61) |
| Percent unemployed | 3.31 (0.74) | 4.96 (1.44) | 3.80 (1.44) | 4.04 (0.91) |
| Percent children in poverty | 11.27 (4.08) | 29.85 (7.02) | 17.47 (4.90) | 19.16 (4.30) |
| Income ratio (80th to 20th percentile) | 4.28 (0.66) | 4.99 (0.78) | 4.15 (0.43) | 4.45 (0.59) |
| Percent households with overcrowding | 2.22 (1.58) | 2.75 (1.96) | 2.02 (1.54) | 2.22 (1.48) |
| Median income | 71557.97<br>(14989.20) | 40188.09<br>(6170.90) | 52465.77<br>(6985.36) | 53390.72<br>(6798.91) |
| **Health Status** |  |  |  |  |
| Years of potential life lost rate | 5755.71 (1127.19) | 10767.91<br>(2061.60) | 7410.08<br>(1550.12) | 8334.47<br>(1203.43) |
| Percent reporting fair/poor health | 13.56 (2.12) | 22.78 (3.62) | 15.11 (2.44) | 17.86 (2.08) |
| Average physically unhealthy<br>days/month | 3.36 (0.37) | 4.70 (0.52) | 3.61 (0.43) | 4.03 (0.33) |
| Percent smoking | 14.04 (2.12) | 20.67 (2.99) | 15.70 (1.81) | 17.93 (2.01) |
| Percent obesity | 27.92 (4.90) | 35.94 (5.03) | 32.06 (4.37) | 33.85 (3.93) |
| Percent physically inactive | 20.61 (3.88) | 31.86 (4.86) | 25.80 (4.02) | 27.72 (3.68) |
| Cardiovascular disease mortality rate | 339.42 (54.33) | 516.66 (91.38) | 397.78 (66.65) | 432.29 (57.85) |
| Cancer mortality rate | 157.83 (20.17) | 202.21 (31.76) | 177.72 (26.85) | 185.79 (18.50) |
| **Healthcare Access** |  |  |  |  |
| Percent uninsured | 8.15 (3.63) | 13.20 (4.77) | 10.36 (5.11) | 10.72 (4.33) |
| Primary care physician rate | 79.86 (41.33) | 41.38 (23.25) | 56.43 (28.58) | 54.56 (28.65) |
| Preventable hospitalization rate | 3912.93 (1238.16) | 5983.81<br>(1953.21) | 4081.10<br>(1573.13) | 4991.30<br>(1028.37) |
| Percent vaccinated | 50.18 (5.64) | 38.93 (7.77) | 40.28 (10.21) | 46.88 (5.49) |
| Hospital beds per capita | 0.00 (0.00) | 0.00 (0.00) | 0.00 (0.00) | 0.00 (0.00) |
| **Environmental and other** |  |  |  |  |
| Social association rate | 9.81 (3.78) | 10.73 (4.17) | 14.07 (5.72) | 10.96 (3.55) |
| Average daily PM2.5 | 8.98 (1.95) | 9.73 (1.33) | 8.06 (1.81) | 10.42 (1.35) |

**Figure 1.**
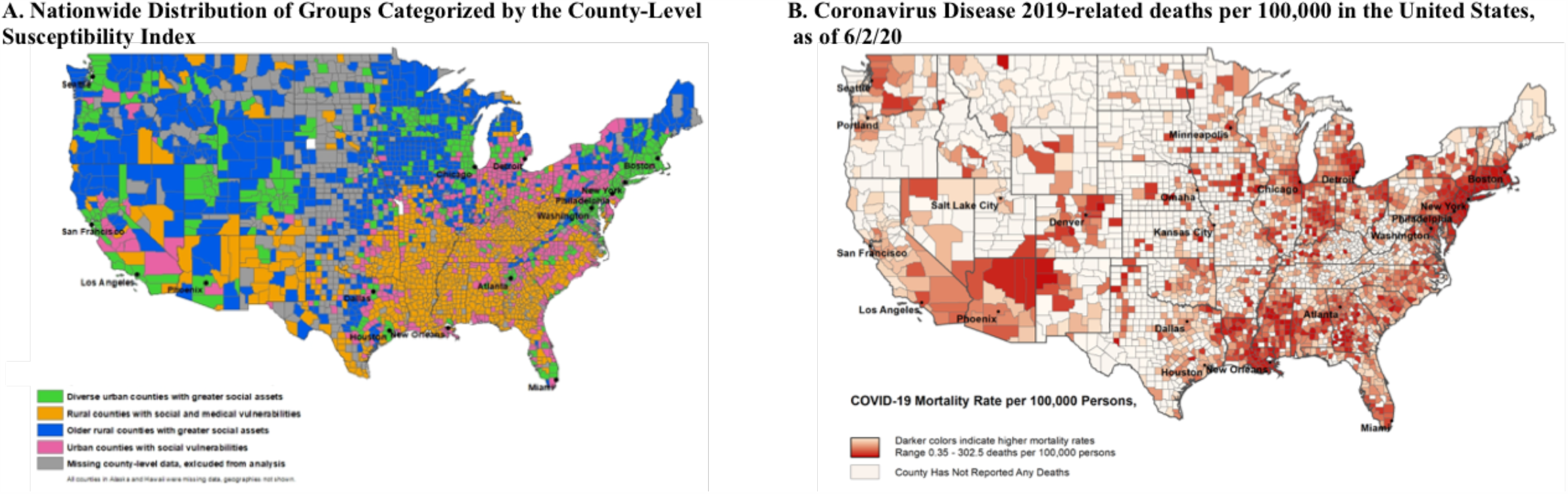
Nationwide County-Level Distribution of Groups Categorized by the County-Level Susceptibility Index (A) and Covid-19 Deaths per 100,000 as of 6/2/20 (B)

Individual county-level metrics included in the latent class modeling were highly correlated with each other (Supplemental Figure 2). For example, favorable social and health assets tracked in counties with lower poverty metrics and higher median income; these counties also had more favorable health metrics such as proportions of smokers, people with obesity, and physically inactive people, as well as fewer years of potential life lost. Conversely, adverse social and health factors and a higher proportion of minority residents were observed in vulnerable counties. These counties also had, on average, lower median income, greater proportion of households with overcrowding, and poorer air pollution rates. Counties within CSI groupings have widespread geographic dispersion with some regional clustering (Figure 2A). For example, rural counties with social and health vulnerabilities arepredominantly, but not exclusively, localized in the southern US.

**Figure 2.**
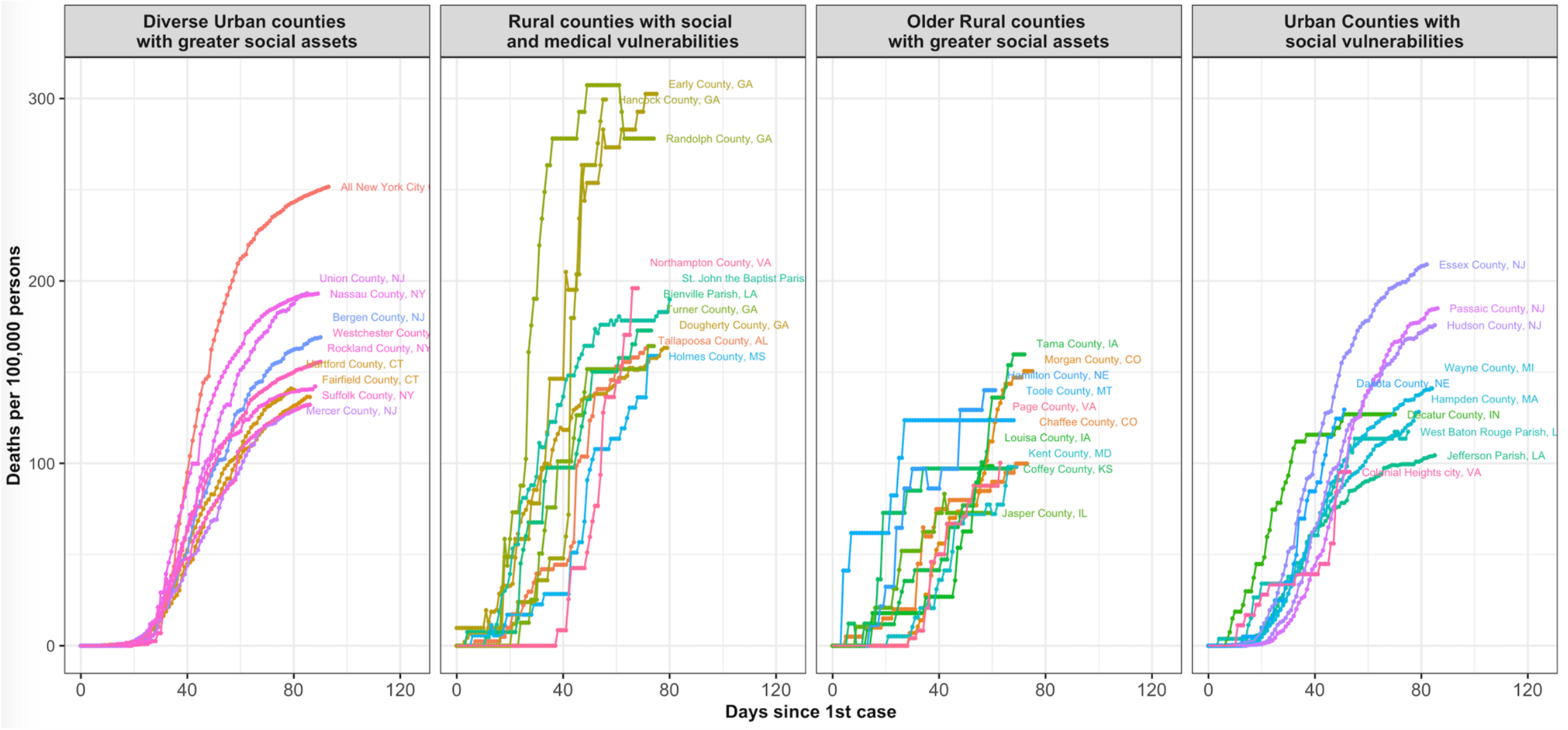
Spatiotemporal Spread of Cumulative Covid-19 Deaths per Capita among the Top Ten Counties* Stratified by County-Level Susceptibility Index. * As of 6/2/20 among counties with reports of 5 or more deaths (N=867 total)

### Covid-19 Related Deaths and Cases Stratified by County-level Susceptibility Index

As of June 2^nd^, 2020, 1735 counties had reported at least 1 confirmed Covid-19 death to be included in the publicly available dataset hosted by the NY Times. Of these, 867 counties had reported at least 5 Covid-19 deaths (Figure 1B) and represented counties in all 4 groups. Within each CSI grouping, we visualized the trajectory of the Covid-19 mortality burden as a function of time since the 5^th^ reported Covid-19 death in the ten counties with the highest cumulative deaths per capita (Figure 2); the burden within CSI groupings is further described in Supplemental Table 3. Qualitatively, CSI groups demonstrate significant heterogeneity with distinct patterns of spatiotemporal spread that are visualized when these same counties are plotted by calendar date (Supplemental Figure 3). While diverse urban communities (e.g. NYC metro area) had earlier spread, the burden of Covid-19 is now emerging in rural counties (e.g. Louisiana and Georgia).

**Figure 3.**
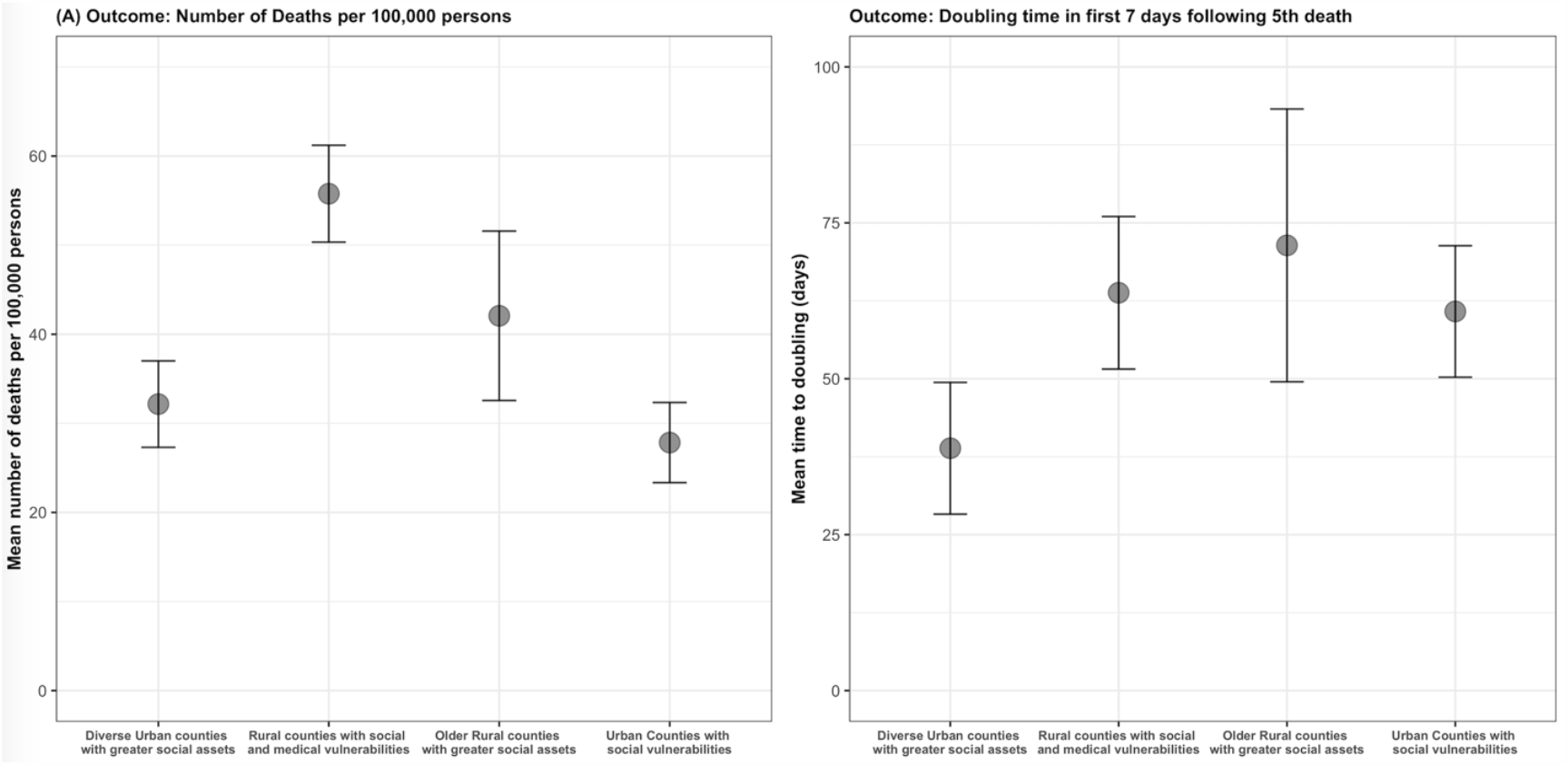
Estimated Marginal Means of (A) Cumulative Covid-19 Deaths per Capita^*^ and (B) Early Mortality Doubling Time^#^, Stratified by County-level Susceptibilty Index. *As of June 2^nd^, 2020 Marginal estimates reported for average time since the first case [(A) 76 days; (B) 40 days] #Early mortality doubling time was calculated in the first 7 days following the 5^th^ death

After adjusting for time since first reported Covid-19 case, Covid-19 deaths per capita varied between CSI groups (Figure 3A, Supplemental Table 4). When comparing counties with 76 days since the 1^st^ case, vulnerable rural counties had more deaths per 100,000 persons compared with diverse urban counties with greater social assets (55.8 [95% CI 50.3-61.2] deaths per 100,000 versus 32.2 [95% CI 27.3-37.0]). Among counties with 40 days since first Covid-19 case report, early mortality doubling times were slower in vulnerable, rural counties compared with diverse urban counties with greater social assets (63.8 days [95% CI 51.6-76.0] versus 38.9 [28.3-49.4], p<0.05; Figure 3B, Supplemental Table 4), but there was significant variability in this distribution within each CSI group (Supplemental Figure 5).

## DISCUSSION

In this study, we described a novel county-level classification scheme based on ecological factors that may be associated with mortality burden related to Covid-19 that identified 4 distinct CSI groupings. This classification scheme leveraged and integrated county-level metrics across multiple key domains (sociodemographic, health-related, and environmental) from multiple data sources to distinguish patterns in underlying county-level vulnerability and resilience. While there was a significant difference in early mortality doubling time related to Covid-19, with faster doubling in diverse urban counties with greater social assets, the burden of Covid-19 deaths per capita is significantly higher in rural counties with social and medical vulnerabilities. The CSI classification scheme identified discordance in the early slope or rapidity of spread within the first week and total loss of life per capita due to Covid-19. We posit this discrimination is likely related to diverse urban counties representing the initial Covid-19 outbreak areas in the US, likely due to the high population density and more frequent international travel in these coastal areas, with a later shift of burden towards the rural and vulnerable Southern counties.

Our findings are consistent with recent data highlighting the “rural mortality penalty” that has emerged in the US since the 1980’s and has been widening in recent years.^24^ This place-based health disadvantage has been demonstrated for all-cause mortality as well as leading causes of noncommunicable disease, such as cardiovascular disease and cancer, ^25,26^ and likely reflects convergence of multiple ecological factors related to social determinants of overall health, such as income, education, and race. As spatial and temporal patterns of Covid-19 remain dynamic, the CSI classification may inform location choices for future policy and intervention strategies to address the burden of future waves, as the CSI scheme reflects differences in underlying vulnerabilities based on social determinants of health in addition to comorbidity burdens and air pollution exposure at a meaningful geographic scale (county-level). This is especially important given greater strain on already limited health resources, such as lower available intensive care unit beds per capita and access to ventilators and hemodialysis machines in rural compared with urban areas.^27,28^

Data about the epidemiologic and clinical characteristics of Covid-19 positive patients to-date include multiple studies highlighting the role of age as a major risk factor for disease severity and outcomes. In the US, the Covid-19-Associated Hospitalization and Surveillance Network (Covid-NET)^29^ analyzed data from 1,482 patients hospitalized with Covid-19 between March 1 and 28, 2020. Highest rates of hospitalization were observed among adults aged ≥65 years and older, consistent with prior studies. While 25% of hospitalized patients in the US were <50 years old, 85% of these younger adults had an underlying condition. The most common underlying comorbidity was obesity (59%) followed by chronic lung disease (36%), and diabetes (20%). Older adults were more likely to have underlying CVD (51%) and hypertension (73%). Approximately 33% of patients were non-Hispanic black and 8% were Hispanic.^12^ Vulnerability as a result of underlying comorbidities may be one important factor in the observed place-based disparities, but does not account for intrinsic systemic and structural racism within our healthcare system that may also be contributing.^30^ Our study incorporates both sociodemographic factors and burden of chronic diseases at the county-level and provides a means to estimate suceptibility and burden of Covid-19 at the population level in hopes of informing equitable resource allocation^31^ and mitigating avoidable health inequalities, especially in counties with limited resources.

Strengths of our study include use of innovative statistical methods in an ecological study leveraging a latent-class modeling approach to county-level categorization that identifies counties with similar profiles of ecological risk factors that may be associated with Covid-19 transmission and mortality. Moreover, to accomplish this task, we harmonized and linked numerous publicly available county-level datasets to create a comprehensive classification scheme that integrates multiple county-level characteristics across multiple domains.

Limitations of our analysis include potential misclassification of Covid-19 mortality data due to under-ascertainment of Covid-19 cases due to variable testing availability and resources across the US. Further, we were not able to adjust for testing per capita at the county level as this information is only available at the state-level. However, our analysis relied on the available comprehensive estimates of cumulative Covid-19 mortality at the county-level and incorporated novel methodologic approaches. Policy changes during the study period (e.g. social distancing, shelter-in-place) were heterogeneous across counties and states and therefore were not adjusted for, but may have biased our results towards the null. Finally, our analysis does not account for individual-level characteristics or predictors of disease severity. However, the current study is ecological. It is not meant to infer causation; rather it is meant to inform large-scale comparisons at the population-level to communicate with local health departments and leaders to inform decisions at the county-level.^32^

The findings from this current study provide unique insights into county-level characteristics that may inform the lack of reserve or resilience at a population-level when challenged by an unexpected stress of a highly infectious communicable disease that carries a significant risk of morbidity and mortality, such as Covid-19.^17^ The CSI classification scheme expands upon prior analyses by developing a simplified, yet comprehensive classification scheme that benefits from the use of an unbiased methodologic approach to include all relevant county-level characteristics across sociodemographic, health, and environmental domains without penalty for high degree of correlations between several metrics. Other tools, such as the social vulnerability index (SVI) and County Health Rankings (CHR) have been applied in ecologic analyses for both health and non-health related outcomes. However, the SVI was created by the CDC to inform responses to natural disasters and therefore relies only on demographic variables from the census and does not include health and healthcare access factors, which are integrally related to community outcomes with Covid-19 exposure.^33^ The CHR was developed in 2010 to stimulate community efforts to improve overall health at the county-level,^34,35^ but only ranks counties within states. Therefore, this current study extends prior reports by developing a national, comprehensive, and data-driven classification scheme to reflect social and health vulnerability for the current Covid-19 pandemic as well as future communicable disease outbreaks on the county-level.

In conclusion, we develop an innovative county-level grouping scheme for counties that can be used to inform equitable allocation of resources in places at-risk of greater burden of Covid-19 morbidity and mortality to mitigate place-based health disadvantage related to Covid-19. It has now been well-described that Covid-19 related deaths are amplifying existing racial and geographic disparities at the county-level.^30^ While widespread surveillance, contact-tracing, and immunity testing are urgently needed and will ultimately be required for long-term recovery on the national level, these findings allow for targeted mobilization at the county-level where the burden of Covid-19 per capita is disproportionately greater in the near-term as the outbreak progresses spatiotemporally across the US.

## METHODS

### Data Sources

We harmonized and linked a comprehensive set of county-level measures from a variety of resources including the U.S. Census Bureau, County Health Rankings (CHR), Centers for Disease Control and Prevention (CDC) Wide-ranging Online Data for Epidemiologic Research (WONDER) mortality database, Area Health Resources Files (AHRF), and United States Department of Agriculture (USDA) across all available counties coded by Federal Information Processing Standard county codes (Supplemental Table 1). For the first part of the analysis, we gathered relevant measures from these sources to describe the counties according to the following categories: demographics, socioeconomic status, health status, healthcare access, and environmental factors, independent of Covid-19 burden. For the second part of the analysis, we queried and linked the above socioecological metrics with county-level Covid-19 data from The New York Times GitHub where information on cases and fatalities is updated daily (https://github.com/nytimes/Covid-19-data, as of June 2^nd^, 2020). All data were deidentified and publicly available, and therefore deemed exempt from Northwestern University’s Institutional Review Board.

### Measures

Counties were described using metrics relating to demographics, socioeconomic status, health status, healthcare access, and environmental factors (Supplemental Table 1). Demographic factors included proportion of racial/ethnic minority and older residents in addition to population density and urbanization status. Socioeconomic metrics included rates of varying educational levels, poverty, and housing. Health status included quality of life and morbidity and mortality rates. Healthcare access reflected insurance, hospital beds per capita, and primary care physician density. Finally, the environmental factor was a measure of air pollution.

The primary outcome was county-specific cumulative deaths per 100,000 persons attributed to Covid-19 as of June 2^nd^, 2020 within all counties reporting 5 or more Covid-19 related deaths. As a secondary outcome, we considered the county-specific Covid-19 mortality doubling time (days) at the beginning of the outbreak, which we defined as within the first seven days following the 5^th^ death reported in a particular county. To reliably estimate these doubling times, we further restricted these analyses to counties who reported five or more cumulative deaths for at least three days as of June 2, 2020. Assuming exponential growth, we calculated county-specific time to Covid-19 related mortality doubling by fitting county-specific linear regression models with time since the first report of 5 or more deaths (days) as the independent variable, and logarithm (base 2) of the cumulative number of deaths as the dependent variable. The inverse of the estimates of the county-specific slopes for time since first report of 5 or more deaths became our secondary outcome.

Due to inconsistencies in the reporting of Covid-19 related deaths, some counties showed decreasing cumulative death counts. To avoid negative estimates of doubling times, we applied the pool adjacent violators algorithm^23^ to the raw county-specific cumulative death counts to ensure they were monotonically increasing prior to fitting the county specific linear regression model described above (N = 34 counties). Counties whose estimated initial Covid-19 related mortality doubling time was 240 days or longer were assigned a value from a single imputation from a uniform distribution with a minimum of 120 and maximum of 365 days (N = 157 counties).

### Statistical Analysis

We used latent class analysis (LCA) to classify US counties into groups based on the included metrics. We included all counties for which we had complete ecological data across county-level metrics that span demographic, socioeconomic status, health status, healthcare access, and environmental factors (N=2,701 counties). Groups of counties were assigned to a county-level susceptibility index (CSI) using model-based predicted posterior probabilities. The LCA model incorporated binary and ordinal variables as-is, and continuous variables were categorized as tertiles before inclusion. To select the optimal number of classes (1-5) for the final LCA model, we imposed the following criteria: (1) qualitative evaluation of distinct predicted classes or groups, (2) ensuring no group had fewer than 5% of counties, and (3) minimizing the Bayesian information criterion. For each of the four final CSI groups, we described county-level characteristics using means, medians, and proportions as appropriate.

The primary analysis, conducted in counties reporting both outcomes (N = 867 counties), consisted of calculating the associations between CSI group assignment and each outcome described above using linear regression models. We fit two sets of models: 1) unadjusted and 2) adjusted for the time since the first confirmed case in the county, to allow comparison between counties at similar points in their outbreak. Point estimates were interpreted in context of their magnitude and Wald-style 95% confidence interval.

We completed two additional analyses to explore the sensitivity of our estimates. First, to understand whether our choice of analytic sample substantially affected the associations between CSI groups and cumulative deaths per 100,000 persons, we repeated the primary analysis within all counties reporting 1 or more COVID-19 related fatality as of June 2, 2020 (N = 1735). Second, we evaluated the influence of our selected starting point for estimating the associations between CSI groups and initial COVID-19-related mortality doubling time. We expanded our analytic sample to include any counties with at least three days of reporting one or more cumulative deaths in the seven days following the first report of one or more deaths in the county (N = 1,701 counties). As described above, we calculated the initial COVID-19-related mortality doubling time beginning at the first report of any COVID-19 related fatality, correcting data discrepancies as needed (987 counties), and repeated the primary analysis. All analyses were completed using R version 3.6.3 or newer.

## Data Availability

All data utilized in this manuscript are publicly available.

## Acknowledgements

The funding sponsor did not contribute to design and conduct of the study, collection, management, analysis, or interpretation of the data or preparation, review, or approval of the manuscript. The authors take responsibility for decision to submit the manuscript for publication and had full access to all the data in the study and take responsibility for the integrity of the data and the accuracy of the data analysis.

